# Quantifying the potential dominance of immune-evading SARS-CoV-2 variants in the United States

**DOI:** 10.1101/2021.05.10.21256996

**Authors:** Pratha Sah, Thomas N. Vilches, Affan Shoukat, Meagan C. Fitzpatrick, Abhishek Pandey, Burton H. Singer, Seyed M. Moghadas, Alison P. Galvani

## Abstract

Recent evidence suggests that some new SARS-CoV-2 variants with spike mutations, such as P.1 (Gamma) and B.1.617.2 (Delta), exhibit partial immune evasion to antibodies generated by natural infection or vaccination. By considering the Gamma and Delta variants in a multi-variant transmission dynamic model, we evaluated the dominance of these variants in the United States (US) despite mounting vaccination coverage and other circulating variants. Our results suggest that while the dominance of the Gamma variant is improbable, the Delta variant would become the most prevalent variant in the US, driving a surge in infections and hospitalizations. Our study highlights the urgency for accelerated vaccination and continued adherence to non-pharmaceutical measures until viral circulation is driven low.

## Introduction

As of June 28 2021, at least seven different COVID-19 vaccines have been rolled out worldwide and more than 200 additional vaccine candidates are in various stages of clinical trials. In the US, more than 338 million doses of vaccines have been administered as of July 18, 2021, and 48% of the population is fully vaccinated. While the daily number of cases has fallen since the apex of the pandemic, the relative prevalence of new variants with a selection advantage−from increased transmissibility, immune escape or both− is on the rise. The B.1.1.7 (Alpha) variant has been the predominant variant in the US since March 2021 until recently and is considered 50% more contagious than the original COVID-19 strain (1) with higher mortality risk (2). The P.1 (Gamma) and B.1.617.2 (Delta) variants are likely more contagious than Alpha.

The three authorized vaccines in the US−produced by Pfizer-BioNTech, Moderna, and Johnson & Johnson−are highly effective for preventing symptomatic and severe disease caused by the originally identified Wuhan-1 strain. Since these vaccines target the spike (S) protein of the original strain, vaccine-mediated neutralizing antibodies may be less protective against newer variants with multiple spike mutations (3–5). Indeed, early laboratory data suggest a significantly lower neutralizing activity for vaccine-induced antibodies against the both B.1.351 (Beta) and Gamma variants (3). Recent studies have also noted lower neutralizing antibody titers for Beta and Gamma in sera from fully vaccinated or previously infected individuals (4, 5). However, the repercussions of reduced neutralizing activity for the protection offered by vaccines remains undetermined.

The increasing prevalence of the Delta variant in the US and the possibility of vaccine escape have raised concerns about the ability of the first generation of COVID-19 vaccines to end the pandemic. The US Centers for Disease Control and Prevention (CDC) guidelines allow fully vaccinated individuals to resume pre-pandemic activities and social interactions (6). Premature relaxation of social distancing measures for all individuals could enhance the selection advantage for immune escape variants, hampering control of the pandemic. Here, we quantified vaccine and immune-mediated selection pressures that could facilitate the dominance of the Gamma or Delta variants in presence of other circulating variants and under the CDC guidelines. We then identified the range of transmissibility and degree of vaccine escape for variants that could lead to a resurgence in cases and hospitalizations in the US.

## Materials and Methods

We extended our previous age-stratified agent-based model of COVID-19 to include the transmission dynamics of the Alpha, Gamma, and Delta variants in addition to the original strain (7). We included these four variants in our model because their prevalence as determined by sequence data surpassed 1% in the US during the evaluated time-period (<u>Trends 2021</u>). The model was parameterized with the population demographics of the US, a contact network accounting for pandemic mobility patterns, and age-specific risks of severe health outcomes due to COVID-19 (SI Dataset 1). We calibrated the transmission probability of the original strain by fitting the model to reported cases in the US per 100,000 population from October 1, 2020, to June 28, 2021. During calibration, we introduced the Alpha variant on December 1, 2020, with a 50% higher transmissibility and 30% higher mortality rate compared to the original strain. The Gamma variant was introduced on January 5, 2021, with 60% higher transmissibility compared to the original strain followed by the Delta variant on March 13 (SI Appendix, Extended Methods).

Vaccination started on December 12, 2020 for 16+ year olds, and May 13, 2021 for 12-15 year olds, following the average weekly rate of doses administered with the Pfizer-BioNTech and Moderna vaccines. The vaccines were modeled as “leaky”, whereby all vaccinated individuals have reduced probability of acquiring infection, probability of developing symptomatic disease if infection occurred, and probability of developing severe disease if symptomatic disease occurred (8). The reductions in these probabilities were drawn from published estimates on vaccine efficacy by variant and by time since vaccination (SI Dataset 3). We adjusted contact patterns in the model to allow fully vaccinated individuals to return to normal pre-pandemic behavior 14 days after the second dose of vaccine, starting from April 3, 2021 (6).

We projected the total hospitalizations caused by different variants between April 1 and December 31, 2021 (Fig.1: A1). Given the lack of data on the selection advantages of the circulating variants, we investigated two key parameters: (i) degree of vaccine escape for the Gamma and Delta variants quantified as a reduction of vaccine efficacy against infection ranging from 0% to 50%, and (ii) the transmissibility of the Delta variant relative to Alpha, which was increased with a multiplicative factor in the range 1 to 1.5. Further details of the model structure and parameterization are provided in SI Appendix, Extended Method. The simulation codes are available at: https://github.com/thomasvilches/multiple_strains.

**Fig. 1.**
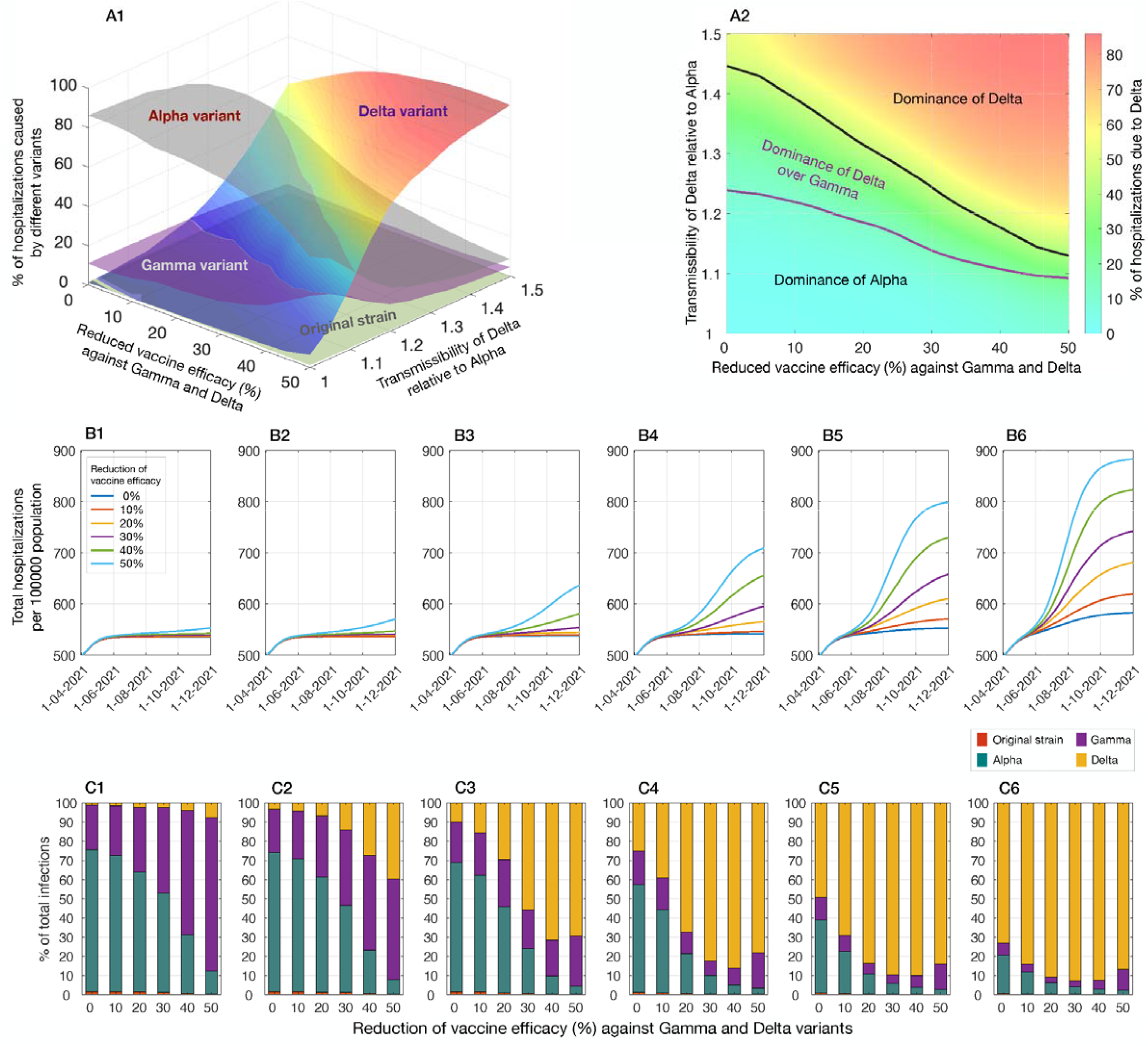
Projected hospitalizations and infections caused by the variants and the original strain betw en April 1 to December 31, 2021. (**A1**) Percentage of hospitalizations due to infections by different variants, with varying transmissibility of Delta relative to Alpha, and reduced efficacy of Pfizer-BioNTech vaccines against Gamma and Delta. (**A2**) Dominance regions in percentage of hospitalizations caused by Alpha, Gamma, and Delta separated by the threshold curves. (**B1-B6**) Total hospitalizations per 100,000 population for relative transmissibility of Delta: 1 (**B1**) 1.1 (**B2**), 1.2 (**B3**), 1.3 (**B4**), 1.4 (**B5**), and 1.5 (**B6**). (**C1-C6**) Percentage of total infections caused by different variants corresponding to the relative transmissibility of Delta in B1-B6.

## Results

We found that the Alpha variant would remain responsible for the majority of hospitalizations between April 1 to December 31, 2021 as long as the increased transmissibility of Delta compared to Alpha remains below 10% (Fig 1, A1, A2). However, the Delta variant would be the most prevalent variant and cause a surge in hospitalizations if its selection advantage, determined by a combination of its transmissibility relative to Alpha and reduced vaccine efficacy exceeds a threshold curve (Fig 1, A2). If vaccine efficacy against the Delta variant is not reduced, we found that the relative transmissibility of Delta would need to be at least 24% and 45% to cause more hospitalizations than Gamma and Alpha, respectively. For example, if Delta is 50% more transmissible than Alpha, we project a resurgence with 15 (95% CrI: 12 − 18) cases per 100,000 population at the peak, causing a total of 89 (95% CrI: 80 − 98) hospitalizations per 100,000 population over the evaluation period, corresponding to approximately 294,000 (95% CrI: 266,000 − 323,000) hospitalizations for the entire US. In this case, the proportion of hospitalizations associated with Alpha, Gamma, and Delta variants are projected to be 39%, 5%, and 55%, respectively.

With vaccine escape capability, Delta can cause more hospitalizations than other variants with an even lower relative transmissibility. For instance, if the reduction of vaccine efficacy is 30%, the Delta variant with a relative transmissibility of 30% would lead to a peak incidence of 28 (95% CrI: 23 − 36) cases per 100,000 population. We project 107 (95% CrI: 96 − 119) total hospitalizations per 100,000 population over the evaluation period, corresponding to approximately 355,000 (95% CrI: 316,000 − 395,000) for the entire US. In this scenario, the proportion of hospitalizations due to Alpha, Gamma, and Delta variants would be 32%, 6%, and 61%, respectively, with only 1% being associated with the original strain. In an extreme scenario with 50% reduction of vaccine efficacy, the proportion of hospitalizations attributed to Delta would be higher than those caused by Alpha and Gamma when the relative transmissibility of Delta exceeds 9% and 14%, respectively.

Our results indicate that an increase in severe disease outcomes due to an immune-evading variant would require a relatively high selection advantage, even when the variant can lead to a resurgence of cases (Fig 1, C1-C6). For example, when vaccine efficacy is reduced by 20%, the Delta variant with a relative transmissibility of 30% would be responsible for the majority of infections over the evaluation period (Fig 2). However, due to high efficacy of vaccines against symptomatic and severe disease, the number of hospitalizations caused by Delta would still remain lower than Alpha during the evaluation period.

**Fig. 2.**
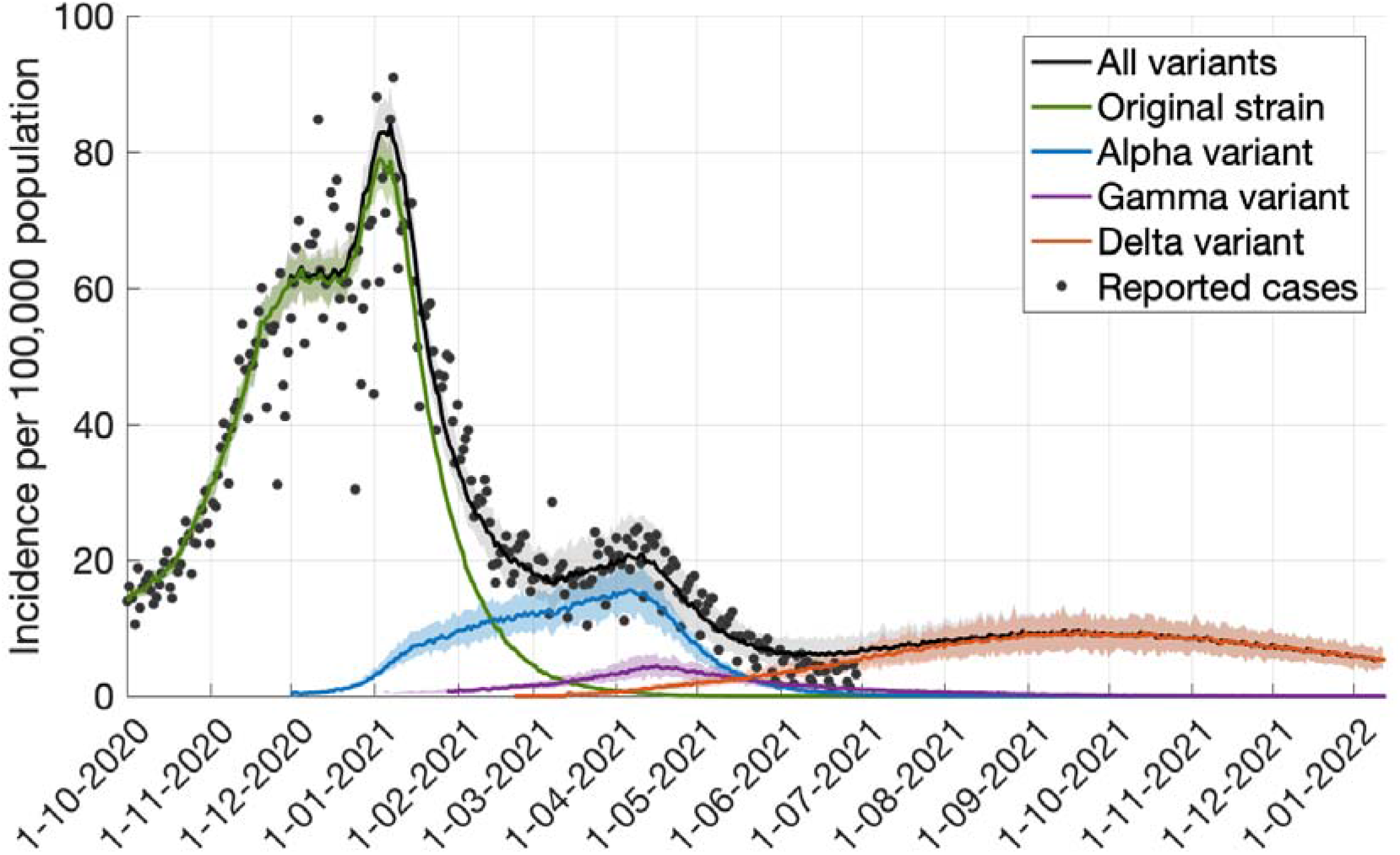
Model fit to COVID-19 case incidence data per 100,000 population in the US, and projected incidence for different variants. The transmissibilities of Alpha and Gamma variants were assumed to be 1.5 and 1.6 relative to the original strain. The transmissibility of Delta was 30% higher than Alpha. Vaccine efficacy against infection with Gamma and Delta was assumed to be reduced by 20% compared to the original strain.

## Discussion

Growing evidence suggests that the Delta variant may be up to 50% more transmissible than the Alpha variant (9), and may be associated with reduced vaccine efficacy (10). Our results suggest that the Delta variant would be the dominant variant in the US, causing a surge in infections and hospitalizations after months of declining incidence. While the dominance of Gamma in terms of infection and hospitalizations appears improbable at the national level in the US, regional flare-ups or congregate outbreaks of this variant may still occur. Corroborating this is local dominance of other variants in California (11) and the low national prevalence but clustered outbreaks of the Gamma variant in Canada (12).

As vaccine rollout proceeds slowly in many countries, the emergence and circulation of new variants will continue to pose challenges to global pandemic control. Selection pressure from naturally-acquired or vaccine-mediated immune responses will inevitably foster the rise of resistant mutations if community transmission remains high. Furthermore, highly transmissible, immune-evading variants such as Delta have the potential to erode the effectiveness of natural and vaccine-elicited immunity, and cause a resurgence of cases, hospitalizations and deaths. The introduction of such variants to the US from countries with limited resources and vast unvaccinated populations will likely continue, highlighting how international efforts to address the global disparities in COVID-19 vaccine distribution are also in the best interest of the US.

Although the emergence of novel variants cannot be completely averted, steps can be taken to mitigate the risks. Reducing community transmission with the available public health tools also lowers the probability of more transmissible variants emerging. While increasing vaccine uptake, adherence to non-pharmaceutical measures should be continued until viral circulation is driven low. Our results further show that despite the evidence of vaccine escape, vaccination remains an effective strategy to combat the pandemic.

## Data Availability

All data are available in the main text or the supplementary materials. The simulation codes are available at: https://github.com/thomasvilches/multiple_strains

## Acknowledgments

APG acknowledges funding from NSF Expeditions grant 1918784, NIH grant 1R01AI151176-01, NSF grant RAPID-2027755 and the Notsew Orm Sands Foundation. SMM was supported by the Canadian Institutes of Health Research [OV4 – 170643, COVID-19 Rapid Research] and NSERC EIDM grant. MCF is grateful for support from NIH grant 5K01AI141576.

