## Supplementary Information Text for "Quantifying the potential dominance of immune-evading SARS-CoV-2 variants in the United States"

*Alison P. Galvani

**This PDF file includes:**

Extended Methods

Legends for Datasets S1 to S3

SI References

**Extended Methods**

**Model structure**

We extended our previous agent-based model of COVID-19 transmission and vaccination (1, 2) to include B.1.1.7 (Alpha), P.1 (Gamma) and B.1.617.2 (Delta) variants of SARS-CoV-2 with different transmissibilities in addition to the original strain. The model implemented the natural history of disease with epidemiological classes for susceptible; latently infected (not yet infectious); asymptomatic (and infectious); pre-symptomatic (and infectious); symptomatic (and infectious) with either mild or severe illness; recovered; and dead. The population was stratified into six age groups of 0 to 4, 5 to 19, 20 to 49, 50 to 64, 65 to 79, and 80+ years (based on demographics of the US population (3) and incorporated age-specific risk of hospitalizations and deaths, contact patterns, and a two-dose vaccination rollout. Daily contacts between individuals were sampled from a negative-binomial distribution parameterized (Table S1) using empirical data on pre-pandemic and pandemic-era interactions (4, 5).

**Transmissibility**

Risk of infection for susceptible individuals depended on their interaction with infectious individuals in the pre-symptomatic, symptomatic, or asymptomatic stages of infection. The transmission probability, which quantifies the risk of infection acquisition for uninfected individuals given a contact with an infectious individual, for the original strain of SARS-CoV-2 was calibrated by fitting the model to case incidence data per 100,000 population in the entire US from October 1, 2020, to June 28, 2021 (6). We chose October 1 as the starting point for our calibration and simulations because it was a time of a relatively low incidence preceding the fall/winter wave in the US and key events investigated in our paper, including launch of the vaccination campaign and emergence of different variants occurred either during or right after the December 2020 - January 2021 surge. During the calibration process (in the presence of only the original strain of SARS-CoV-2), we used a 50% lower rate of contacts (compared to pre-pandemic normal behaviour) and determined the transmission probability of 0.109 that minimized the difference between mean cumulative incidence predicted by the model and cumulative reported cases. This transmission probability obtained during the calibration corresponds to an effective reproduction number of 1.17 in early October 2020 (7). After the calibration, the transmission probability of the original strain remained fixed and the age-specific contact rates were adjusted throughout the simulations to implicitly account for the change in various NPIs implemented in the US. Specifically, the age-specific contact rates for all individuals were increased by 6% over 6 days from October 17, 2020, decreased by 6% over 6 days from November 20, increased by 9% over 9 days from December 20, 2020 and decreased by 22% over 16 days from January 3, 2021, followed by the return of fully vaccinated individuals to normal behaviour from April 3, 2021.

We introduced the Alpha variant on December 1, 2020 (12 days prior to the start of vaccination in the US) (8) with a 50% higher transmissibility and 64% higher risk of death compared to the original strain (9–11). We then introduced the Gamma variant on January 5 (12) with a transmissibility that is increased by a multiplicative factor of 1.6 relative to the original strain (13). Finally, the Delta variant (B.1.617.2) was introduced in the model on March 13, 2021, when the first case was identified in the US (14). The transmissibility of this variant was varied with a multiplicative factor from 1 to 1.5 relative to the B.1.1.7 variant (15).

**Disease dynamics**

We parameterized the infectivity of asymptomatic, mild symptomatic, and severe symptomatic individuals to be 26%, 44%, and 89% relative to the pre-symptomatic stage (16–18). We assumed that these relative infectivities remained the same for all variants in the model. The incubation period was sampled from a log-normal distribution with a mean of 5.2 days (19), and parameters of 1.434 (shape) and 0.661 (scale). An age-dependent proportion of infected individuals progressed to a pre-symptomatic stage with a mean duration of 2.3 days, sampled from a Gamma distribution with parameters of 1.058 (shape) and 2.17 (scale) (17, 20). Pre-symptomatic cases developed symptomatic disease with a mean duration of 3.2 days, which was also sampled from a Gamma distribution with parameters of 2.77 (shape) and 1.1563 (scale) (21, 22). The remaining proportion of infected individuals experienced asymptomatic infection until recovery, with a mean infectious period of 5 days sampled from a Gamma distribution with parameters of 5 (shape) and 1 (scale) (21, 22).

Recent studies indicate that antibodies from prior infection with other variants of SARS-CoV-2 may have reduced neutralizing activity against Beta, Gamma and Delta variants (23–26). We therefore assumed that similar to Beta, both Gamma and Delta variants evade naturally acquired immunity by an average of 21% (95% CI: 11-36%) (27, 28). This evasion rate was implemented as a reduction of immune protection for individuals recovered from the original strain or the Alpha variant, corresponding to a reduced average transmission probability of 0.0229 (21% of the original 0.109 transmission probability used in the simulations) for reinfection by the Gamma or Delta variant. We further assumed that recovery from infection by the Gamma or Delta variant provides protection against all variants in the model, preventing reinfection for at least one year.

**Infection outcomes**

We assumed that asymptomatic and mild symptomatic cases recover from infection without hospitalization. A proportion of those with severe disease were hospitalized within 2-5 days of symptom onset (29, 30) and were therefore removed from the transmission chain. We also assumed that all symptomatic cases who were not hospitalized self-isolated within 24 hours of symptom onset, and reduced their number of daily contacts by an additional 72% (Table S1). Intensive care unit (ICU) and non-ICU hospitalization rates were parameterized (Table S2) by clinical and epidemiological data stratified by age and comorbidities (31–33).

**Vaccination**

We implemented a two-dose vaccination campaign with a sequential prioritization of: (i) healthcare workers (5% of the total population) (34), adults with comorbidities, and those aged 65 and older; and (ii) other individuals aged 16-64 (35, 36). Based on vaccine uptake data, we assigned 60% probability of vaccination for individuals aged 40-64 years and 40% vaccination probability for individuals aged 16-39 years (37). The minimum age-eligibility for vaccination was 16 years before May 13, 2021 after which children aged 12 to 15 years became eligible for vaccination. We used reported daily vaccine doses administered since the start of vaccination to parameterize a rolling 7-day average of vaccine distribution per 100,000 population (38).

We specified Pfizer-BioNTech vaccines with an interval of 21 days between the first and second doses (39). We parameterized the model with published estimates of vaccine efficacy following each dose of Pfizer-BioNTech vaccines against infection, symptomatic disease, and severe disease caused by the original strain (1, 40). These efficacies, reported in Table S3, were implemented in the model as a reduction in probability of acquiring infection, probability of developing symptomatic disease if infection occurred, and probability of developing severe disease if symptomatic disease occurred. We used published estimates for Pfizer-BioNTech vaccines against infection and severe disease with the Alpha and Gamma (assumed to be similar to those reported for Beta). For scenarios of vaccine escape (due to reduced neutralizing activities), we also varied a multiplicative factor for reduction of vaccine efficacy against infection in the range 0-50% for both Gamma and Delta variants (23, 24, 26, 41, 42).

**Model implementation**

We simulated the model with a population of 100,000 individuals from October 1, 2020 to December 31, 2021 assuming a 10% pre-existing immunity generated by the original strain prior to October 2020 (43, 44). To incorporate the age distribution of pre-existing immunity in the population, we ran the model with only the original strain in the absence of vaccination and determined the infection rates in different age groups when the overall attack rate reached 10%. The distribution of this immunity was used to parameterize the initial population at the start of simulations. Vaccination was initiated on December 12, and rolled out as a two-dose strategy with the age-specific distribution of first and second doses (37).

On April 2, the guidelines by the US Centers for Disease Control and Prevention indicated a minimal risk for fully vaccinated individuals to travel and engage in certain social activities while taking COVID-19 precautions (45). We therefore allowed vaccinated individuals to return to normal pre-pandemic behaviour 14 days after the second dose of vaccine from April 3, 2021. The model was implemented in Julia, which is an open-source, high-performance, dynamic programming language that allows rapid analysis of computationally intensive problems, such as agent-based modelling. The simulation codes are available at:

<https://github.com/thomasvilches/multiple_strains>

**SI Dataset 1:** Mixing patterns and the daily number of contacts derived from empirical observations. Daily numbers of contacts were sampled from negative binomial distributions for different scenarios. Parametrization of the Negative Binomial distributions can be derived from the given mean and standard deviations. NPIs stands for Non-Pharmaceutical Interventions.

**SI Dataset 2:** Model parameters associated with hospitalization of severe cases.

**SI Dataset 3:** Estimated vaccine efficacies (%) from published studies.
